# Classifying Alzheimer’s Disease and Dementia Patients Using Non-invasive EEG Biomarkers

**DOI:** 10.1101/2024.10.03.24314841

**Authors:** Wali Hasan, Sheraz Khan, Abbas Sohrabpour

**Affiliations:** 11^th^ Grade Student, Lake Highland Preparatory High School, 901 Highland Ave, Orlando, FL 32803; Athinoula A. Martinos Center for Biomedical Imaging (MGH), Boston, MA 02129

## Abstract

Researchers are currently exploring methods to detect early-stage Alzheimer’s Disease (AD) and other forms of dementia such as frontotemporal dementia (FTD), especially through non-invasive biomarkers, i.e., measurements that reflect biological processes. This paper utilizes a dataset of electroencephalogram (EEG) recordings, a noninvasive biomarker, to distinguish individuals with Alzheimer’s or frontotemporal dementia from healthy control subjects. This paper explores the usage of machine learning methods to more accurately predict the cognitive status of patients from these non-invasive EEG Biomarkers. We found that AD patients could be easily separated from healthy controls based on their EEG features using simple linear classifiers with an accuracy of 77% and the AD, FTD, and healthy controls with an accuracy of around 57% (randomly selecting the right class is about one third or 33% in the 3-way classification).

## 1 Introduction

Approximately fourteen million people are projected to develop Alzheimer’s Disease (AD) by 2060, three times the current Alzheimer’s population size [MXG^+^19]. Alzheimer’s Disease is a progressive brain disorder that destroys brain cells and synapses, leading to a decline in cognitive ability that eventually results in dementia. Additionally, Alzheimer’s disease is highly associated with and, due to its degenerative properties, may even cause mental health issues such as depression, apathy, aggression, and psychosis [LHT^+^14]. While Alzheimer’s disease affects the whole brain, frontotemporal dementia (FTD), the second leading cause of early-onset dementia, affects the frontal regions of the brain [BSM15]. Currently, there is increasing interest in biomarkers for the early detection of these neurodegenerative diseases. This paper explores electroencephalogram (EEG) recordings, measurements of electrical activity produced by the brain, as a noninvasive biomarker in detecting Alzheimer’s disease and frontotemporal dementia using machine learning.

Previous studies have investigated the efficacy of EEG oscillations’ frequency content or spectral power [PVLGM^+^21]. We follow on these works by considering EEG features derived from the power spectrum density (PSD) as features to classify AD vs. other healthy or pathological sub-populations. We employ a few well-known classifiers to achieve this goal. Such ideas and works can one day lead to the early detection of AD even before major cognitive decline has occurred. The *Codes* developed for data analysis and figure production are available from https://github.com/walihas/walihas/blob/main/Classifying_Alzheimer's_Disease_and_Dementia_Patients_Using_Non-invasive_EEG_Biomarkers.ipynb

## 2 Methods

### 2.1 Dataset Description

In order to classify subjects based on pathological conditions into the AD group, healthy control group (CN), and the FTD group from their EEG recordings, a publicly available dataset [MTA^+^24] containing EEG recordings of 36 Alzheimer’s patients, 23 frontotemporal dementia patients, and 29 healthy age-matched subjects was selected. The EEG recordings contained 19 scalp electrodes (following the regular 10-20 system). Each EEG recording was performed with the subject sitting with their eyes closed. The sampling rate was 500 Hz. The dataset was already pre-processed using a Butter-worth band-pass filter with a frequency range of 0.5 to 45 Hz, an artifact subspace reconstruction (ASR) routine [KJ16] that rejected large amplitude artifacts, and ICA that automatically classified and rejected eye and muscle (jaw, swallowing, etc.) artifacts [MTA^+^23].

### 2.2 Feature Extraction

Due to its reflection of brain processes, the relative power spectral density (the frequency of appearance or strength) of typical frequency bands may be an important biomarker for cognitive weakening such as that exhibited in Alzheimer’s disease and dementia. Five frequency bands are particularly significant. These are *δ* (0.5 - 4 Hz); *θ* (4 - 8 Hz); *α* (8 - 13 Hz); *β* (13 - 25 Hz); and *γ* (25 - 45 Hz).

To calculate the relative PSD of each of these bands, EEG recordings were epoched into six-second windows. Then, the PSD of each band was calculated using the Welch method, which uses Fourier transforms to average over windowed power estimates. For each subject, the PSD of each band was averaged per channel. The relative PSD was calculated by dividing the power of each PSD band by the total sum of powers in the whole frequency range (0.5 - 45 Hz). This culminated in seven relative power spectral density features: Average power in *δ, θ, α, β*, and *γ* frequency bands as well as Alpha-to-Theta 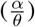 and Beta-to-Theta power ratios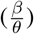.

The average values of each of these features were taken for each group (CN, AD, FTD) and were then plotted on topographical maps (topo-maps) across the scalp. We used the NeuroDSP Toolbox for these analyses [CDGV19]. MNE Python was also used for importing and handling Data[GLL^+^13].

### 2.3 Statistical Tests

The Mann-Whitney U Test was used to derive the p-value (the probability that two distributions are similar) between the spectral power bands of each population (CN, AD, FTD) over all channels.

### 2.4 Machine Learning Classification

Both binary (between AD and CN groups) and multiclass (3-way) classifications (between all subpopulations) were used. We employed the SciKit-Learn toolbox to implement these classifications and algorithms [PVG^+^11]. A pipeline with the following steps was utilized within a nested fold to train and score the machine learning model: a standardized scaler that normalizes the features so that no one feature over-determines the decision-making process; a principle component analysis (PCA) that reduces the dimensions of the extracted features of the dataset to reduce redundancy and ultimately maximize the separability of features; and classifiers such as the linear discriminating analysis (LDA), linear kernel support vector machine (Linear SVC), and a radial basis function (RBF) kernel support vector machine that utilizes features from the dataset to identify the classes. Linear classification essentially works by taking the dot-product of a trained weight vector *w* and feature vector *x* (the average relative PSDs and power ratios, in this paper) [Equation 1].

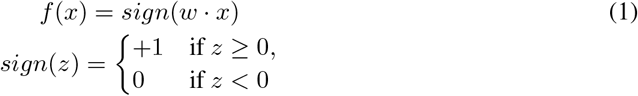

For instance, in a two-class classifier, the linear decision line splits the feature vector space into two half-areas: the positive side, which is on the same side as the normal vector, and the negative side, which is on the opposite side. In contrast, the RBF kernel reaches classification by measuring the similarity in distance between data points in high dimensions. In other words, the closer data points gather, the more likely they belong to the same class. Then, a nested fold cross-validation strategy was used to train the data and to prevent over-fitting. In this strategy, different combinations of train and test splits are used. First, a grid search cross-validation is used to maximize the score by fitting the model to each training set and then optimizing its hyperparameters. Then, a cross-validate score method is used to average test scores over different data splits. The performance of these algorithms was analyzed with statistical metrics, specifically, accuracy, recall, precision, F1 score, and balanced accuracy. Accuracy measures the proportion of total correct predictions among all predictions made. Recall measures the proportion of all true positives among all actual positive instances. Precision measures the proportion of all true positives among all positives predicted by the model. F1 combines precision and recall to measure the accuracy of an individual test. Balanced accuracy is the arithmetic mean of the recall score and the true negative rate, the proportion of true negatives among all actual negative instances.

## 3 Results

### 3.1 Results of the Average Band Evaluation

The graphs of the average power in *θ*, Alpha/Theta 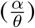, and Beta/Theta power ratios 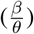 among the three populations as displayed in Figure 1, indicate separability between the features and therefore the classes.

**Figure 1.**
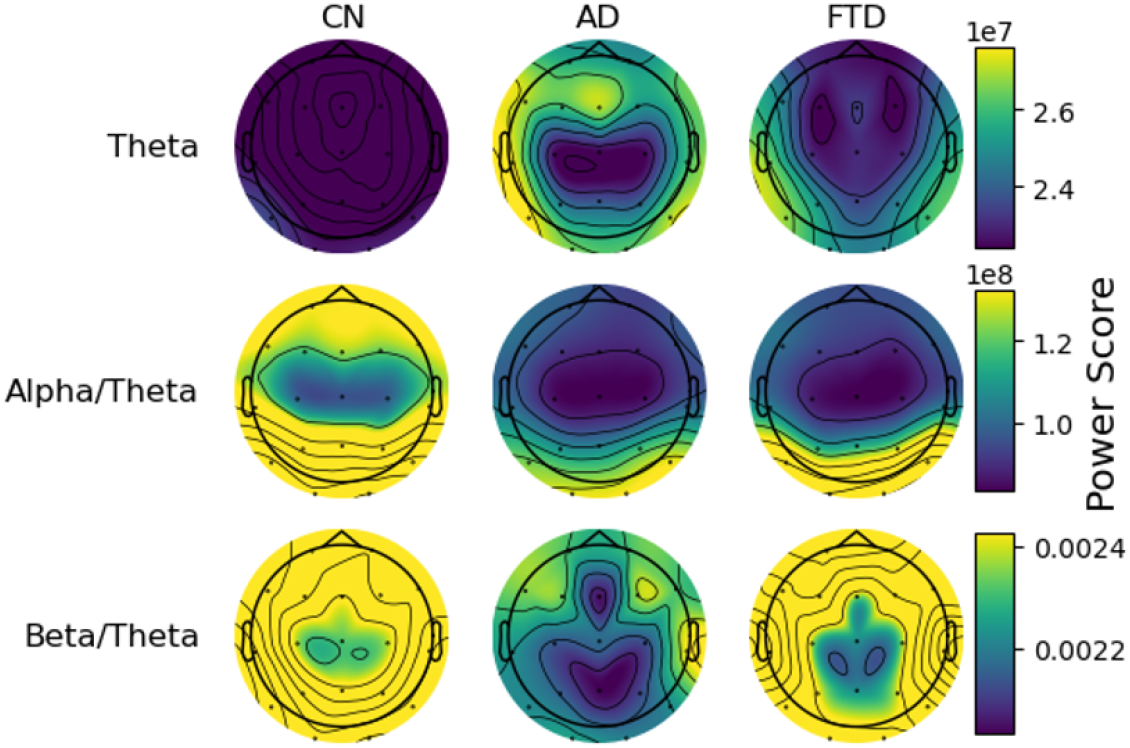
Topographical maps of the average power spectral density for control, Alzheimer’s disease, and dementia groups among several different features: *θ*, Alpha/Theta 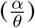, and Beta/Theta 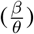.

Although there are notable differences between each population, the starkest differences occur along the *θ, α, β*, Alpha/Theta 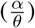 and Beta/Theta 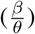 power features. The Alzheimer’s population shows overall more expression of *θ*, less frontal cortex expression of *α*, less posterior expression of *β*, less overall and specifically frontal expression of Alpha/Theta 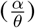, and less overall expression of Beta/Theta 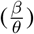. This indicates the possible separation of these populations based on these features.

### 3.2 Results of the Statistical Tests

We found that the *δ, θ, α, β*, Alpha/Theta 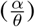, and Beta/Theta 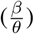 features were significantly different (*p <* 0.05) between CN and AD groups and *θ, β, γ*, Alpha/Theta 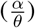, and Beta/Theta 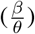 were significantly different between CN and FTD groups, suggesting that these features are appropriate to be used for classification.

### 3.3 Results of Classification

The different performance metrics for each learning model (LDA, SVM with the linear kernel, and SVM with the RBF kernel) indicate the separability of the populations. The results of the CN-AD classification are in Table 1 and the results of the CN-AD-FTD classification are in Table 2. The results indicate that Alzheimer’s classification based on EEG recordings is incredibly plausible as each performance metric is above the chance level indicating that the model can distinguish the classes using EEG features.

**Table 1:** Performance of CN-AD Classification (Mean *±* STD)

|  | Accuracy | Recall | Precision | F1 Score | Balanced Accuracy |
| --- | --- | --- | --- | --- | --- |
| LDA | 77.32%±9.93% | 69.20%±18.78% | 80.32%±15.78% | 72.16%±13.80% | 76.61%±10.24% |
| Linear SVC | 76.12%±10.63% | 72.99%±17.17% | 75.67%±15.62% | 72.65%±13.03% | 75.88%±10.65 |
| SVM (RBF) | 72.80%±10.75% | 70.53%±17.48% | 71.40%±15.18% | 69.29%±12.52% | 72.67%±10.75% |

**Table 2:** Performance of CN-AD-FTD Classification (Mean *±* STD)

|  | Accuracy | Recall | Precision | F1 Score | Balanced Accuracy |
| --- | --- | --- | --- | --- | --- |
| LDA | 55.69%±9.14% | 55.69%±9.14% | 55.69%±9.14% | 55.69%±9.14% | 52.00%±9.02% |
| Linear SVC | 57.54%±9.89% | 57.54%±9.89% | 57.54±9.89% | 57.54%±9.89% | 54.51%±9.66% |
| SVM (RBF) | 51.35%±9.92% | 51.35%±9.92% | 51.35%±9.92% | 51.35%±9.92% | 48.12%±9.56% |

## 4 Discussion

In this paper, we present a non-invasive and inexpensive computational model that can assist in the early detection of Alzheimer’s Disease and frontotemporal dementia based on EEG biomarkers. Our results show that each performance score was above the chance level for each experiment (50% for the CN-AD classification and 33% for the AD-CN-FTD classification) which means that our models are more likely to predict a subject’s class correctly than assigning equal probability for the subject to be any of those classes.

### Limitations

Dementia biomarkers change throughout the different stages of AD and FTD (leading to inaccurate predictions in the model) [JH13] even though our results indicate that differentiating between CN, AD, and FTD groups based on EEG is possible. This could be a source of error. However, with the advent of more complex Machine Learning algorithms such as neural networks, these results could improve further. Based on our experience here, one has to be wary of overfitting the data when using models with more parameters. While we used nested cross folds, as is common in the field, to avoid this problem as much as possible, the limited number of subjects, i.e., a small dataset, makes it difficult to avoid overfitting altogether. In the future, we may be able to better predict these sub-populations by possibly enlarging the database to include patients in many different stages of AD to reflect the changes in biomarkers and/or including other features such as Magnetoencephalography (MEG) to provide the model more information for classification. Furthermore, data should be collected at different institutions and with different devices to ensure that hardware differences do not impede the algorithms’ learning.

### Broader impacts

Overall, because AD and other forms of dementia are projected to affect a large population in the future, EEG represents a beneficial method that can assist medical researchers and physicians in the early diagnosis of different types of dementia. This may not only allow the early prevention of cognitive decline but also the prevention of its associated mental health issues such as depression, a rapidly increasing problem in modern society [LHT^+^14].

## Data Availability

The authors would like to thank the OpenNEURO platform and Miltiadous et al. for making this useful dataset publicly available, online.

https://openneuro.org/datasets/ds004504/versions/1.0.2

## Acknowledgments and Disclosure of Funding

The authors would like to thank the OpenNEURO platform and Miltiadous et al. for making this useful dataset publicly available, online. More information about the implementation details and the codes used for analyzing the data in this paper can be found at: https://github.com/walihas/walihas/blob/main/Classifying_Alzheimer's_Disease_and_Dementia_Patients_Using_Non-invasive_EEG_Biomarkers.ipynb.

